# A novel methodology to measure waiting times for community-based specialist care in a public healthcare system

**DOI:** 10.1101/19013367

**Authors:** Rachel Wilf Miron, Ilya Novikov, Arnona Ziv, Avishai Mandelbaum, Yaacov Ritov, Osnat Luxenburg

## Abstract

**Background:** Monitoring of waiting time (WT) in healthcare systems is essential, since long WT are associated with adverse health outcomes, reduced patient satisfaction and increased private financing.

**Objective:** To develop a methodology for routine national monitoring of WT for community-based non-urgent specialist appointments, in a public healthcare system.

**Design:** Observational historical study using data from computerized appointment scheduling systems of all Health Maintenance Organizations (HMOs) in Israel.

**Data sources:** Data included available appointments for community-based specialists and actual number of visits.

The first 50 available appointments from each specialist appointment book were collected throughout December 2018. Five most frequently visited specialties - orthopedics, ophthalmology, gynecology, dermatology and otolaryngology - were included.

**Data collection/extraction approach:** WT offered to HMO members (OWT) were calculated for two scenarios: “specific” physician and “any” physician in the clinic’s geographical region. Distribution of OWT was calculated separately for each specialty and geographical region, combined to create the nationwide distribution, and expressed as mean, standard deviation and percentiles.

**Principal findings:** Estimated national median OWT for “specific” physician, ranged from 6 days (ophthalmology) to 13 days (dermatology), with large variation between geographic regions. OWT for “any” physician were 28-50% shorter than for “specific” physician.

**Conclusions:** This novel method offers a solution for ongoing national WT measurement, using computerized scheduling systems. It integrates patient preferences for physician choice and allows identification of differences between specialties and regions, setting the ground for interventions to strengthen public healthcare systems.

## Introduction

In 2001, the Institute of Medicine report identified timeliness as one of the fundamental properties of high quality healthcare.[1] The amount of time that a patient needs to wait for a clinician appointment or a treatment, is a key indicator of overall system performance. However, timeliness is still the least studied and least understood dimensions of quality care.[2]

Long waiting times (WT) may negatively affect health outcomes [3, 4] and patient experience with clinical care, including perceived quality of care.[5] Canadian patients waiting for a consultation with a medical specialist for a new medical condition reported worry, stress, anxiety, deterioration of health and loss of work.[6] Longer WT in publicly-funded health systems could increase purchase of private insurance [7] and increase the national health expenditure.

Controlling WT is a major policy concern in publicly funded health systems across OECD countries. The majority of OECD countries monitor and publish national WT statistics and have some form of WT guarantee or target of maximal WT [8]. Most countries measure WT for elective procedures, based on administrative data.[9] In Canada, WT for visits to specialists and for diagnostic and surgical procedures are measured by physician surveys. These surveys measure and report two consecutive WT segments: From referral by a general practitioner to consultation with a specialist, and from the consultation with a specialist to the point at which the patient receives treatment.[10] In the USA, WT for primary and specialist care is measured by “secret shoppers” who describe a non-urgent clinical scenario and ask for the first available appointment.[11] The Veteran Affairs (VA) health administration measures and publicly reports WT across primary and specialty care. WT is calculated as the difference (in days) between the day the veteran requested an appointment and the date of the appointment, representing actual WT in a retrospective approach.[12]

In Israel, a national health insurance system provides citizens with universal coverage and guarantees all citizens the right to health services “at a reasonable time, distance and quality”.[13] Four competing Health Maintenance Organizations (HMOs) provide their members with access to a statutory benefits package. Most specialized ambulatory care is provided in community settings.[14] A “named” or chosen physician is usually recommended by the HMOs and is also the preference of members themselves. All Israeli HMOs have Computerized Appointment Scheduling Systems (CASS). The proportion of physicians that utilize the CASS differs among HMOs, geographical regions and medical specialties, averaging 85% (range 65% to 97%).

Long WTs for consultations and surgical procedures and progressive increase of private financing were the focus of public concern in Israel in recent years: In a public survey carried out in 2012, 28% of respondents reported waiting more than one month for an appointment with a specialist, a period twice as long as the 2 weeks that Israeli health consumers perceive as “acceptable”.[15] Respondents with higher education or those living in central Israel had a higher chance of visiting a community-based specialist.[15] The above mentioned issues of access and equity were among the reasons for creating the Advisory Committee for Strengthening the Public Health System, headed by the Minister of Health. In 2014 the committee published its recommendations, including the recommendation to develop a national measurement system and to make this information available to the public, to set WT targets and use financial incentives in order to encourage providers to achieve those targets.[16] Between 2012 and 2018, the proportion of respondents who visited a specialist in the public system and waited more than a month for their appointment, increased from 28% to 33%; one in three respondents reported seeing a specialist in the private sector, with “desire to reduce WT” quoted as the leading reason for shifting from the public to the private health system.[17]

However, attempts to develop national indices for WT have encountered challenges, mainly due to different information technology (IT) systems, resulting in different measurement approaches among the Israeli healthcare providers. At the time of the study, one HMO had implemented retrospective measurement of WT, while the other three HMOs measure the first or second available appointments offered to their members, in a prospective approach.

In recent years, the issue of significant variation in access to specialist care among specialties and between geographic areas, and its deleterious effects on equity and efficiency, led the Ministry of Health (MOH) to accelerate efforts geared towards the creation of a national WT measurement scheme.

Comparison of methodologies for measurement of WT demonstrates substantial differences among countries. The measurement approach can be retrospective, like the current system at the VA;[12] prospective, i.e. time to the first available appointment, as in a survey of 15 US cities [11] or to the third available appointment, which is used for example as an access measure at Cincinnati Children’s Hospital;[18] or a combined measure. New Zealand, for example, measures time to first specialist appointment – in a prospective approach, as well as retrospective collection of patient flow data.[19] Routine monitoring should be based on operational databases, which are preferred over surveys. Retrospective measures permit estimation of Actual Waiting Time (AWT) if databases include the time of the first attempt to set an appointment and the time of the corresponding actual visit. Currently no Israeli HMO’s database includes all the necessary information. Distribution of WT to first, third or fifth available appointment cannot represent the real distribution of the corresponding WT offered by the HMOs to their members. The three first available appointments may underestimate WT because of cancellations and other last minute changes in appointment schedules. On the other hand, many physicians see less than 100 patients per month. For such physicians, in a steady state (i.e. supply equals demand), no more than three patients on average look for an appointment each day, therefore using fourth or fifth appointment would overestimate OWT. Such considerations led us to develop a different approach, utilizing the exact number of available appointments, representing the expected number of patients looking daily for an available appointment, for each of the physicians.

The first step towards the establishment of a national OWT measurement was directed towards community-based specialty care. It was decided to focus at the first stage on the five most frequent specialties (orthopedics, ophthalmology, otolaryngology, dermatology and obstetrics & gynecology) which comprise 66-70% of community-based encounters with specialists [16, 17] and do not require a physician referral. Information about available appointments for non-urgent care is available to HMO members for self-scheduling on the internet sites or with the help of staff members at designated Call Centers. Appointments for urgent health conditions are provided by the HMOs in diverse ways and are not transparent to the public as available appointments.

The study was designed to develop a methodology for national measurement of WT for non-urgent community-based appointments with specialist physicians in the public healthcare system.

## Methods

### Design and setting

This is an observational historical study using administrative data from all four HMOs in Israel, covering the whole population (except for those in active military service). Institutional review board approval was not required since patient records were not accessed, but rather routine data on next available appointments from HMOs was used. After studying the existing measurement approaches and computerized infrastructures of HMOs, it was decided to adopt a prospective approach based on the HMO’s computerized appointment scheduling systems (CASS) for regular, non-urgent care. All 6,040, community-based physician practices in the following specialties - orthopedics, ophthalmology, obstetrics and gynecology, dermatology and otolaryngology, were included.

A unified data structure was defined to guide data extraction from the HMO operational systems. During 31 consecutive days in December 2018, the first 50 available appointments were extracted daily from the CASS of each physician practice (a specialist can work in more than one practice, each with a separate appointment schedule). This process produced a total of 1,017,478 available appointments for analysis. Concomitantly, the number of actual patient visits at each physician’s practice during the same time period (31 days) was collected.

### The measurement approach

A multidisciplinary team of experts in medicine, computer sciences, statistics, mathematics, and queuing theory, participated in the methodological “journey”. An algorithm was developed, in order to measure WT for available appointments, offered by HMOs to their members for non-urgent care (offered wait time; OWT). The algorithm was designed for two distinct scenarios of patient preferences: a) appointment with a “specific” or “named” physician – when the patient chooses a physician and tries to schedule the first available appointment for the chosen specialist, and b) appointment with “any” physician – when the patient tries to schedule the first available appointment with any physician in a determined specialty and a determined region. Since patients look for available appointments within a reasonable distance from their place of residence, analysis was carried out both at town level and at the wider level of “natural” area (the smallest official classification of the country, consisting of 52 geographical areas).

The following assumptions were utilized to estimate the distribution of OWT: 1) A “steady state” assumption which states that supply equals demand. In other words, the number of actual visits that took place at a particular physician’s practice in a given period of time (supply) is equal to the number of patients who look for available appointments for that physician in the same period of time (demand). Since we did not have demand data, demand was estimated from supply. 2) The daily demand, or the daily number of patients who look for available appointments at a specialist practice, is identical for all the days in that period and is equal to the daily average of actual number of visits that took place at this physician’s practice in this period of time. These assumptions are further examined in the discussion.

### Development of the algorithm

The first 50 appointments available on each day of the study period (“execution date”) were collected from each of the 6,040 appointment schedules. The decision to collect this number of the first available appointments was based on all the assumptions mentioned above and on the maximal number of patients admitted per day across 99.9 percent of all appointment books. On each execution date, WT for all available appointments were calculated as the difference, in days, between the available appointment date and the execution date. This first step of the algorithm is illustrated in Figure 1. For convenience of the illustration, it presents only the first 10 available appointments in one physician’s practice appointment book, for 3 selected execution dates in December 2018. The calculated results are presented for each available appointment date in the book, for example: On 2^nd^ December, WT to the first available appointment (which is on 6^th^ December) is 4 days, while WT to the sixth available appointment, on 10^th^ December, is 8 days.

**Figure 1:**
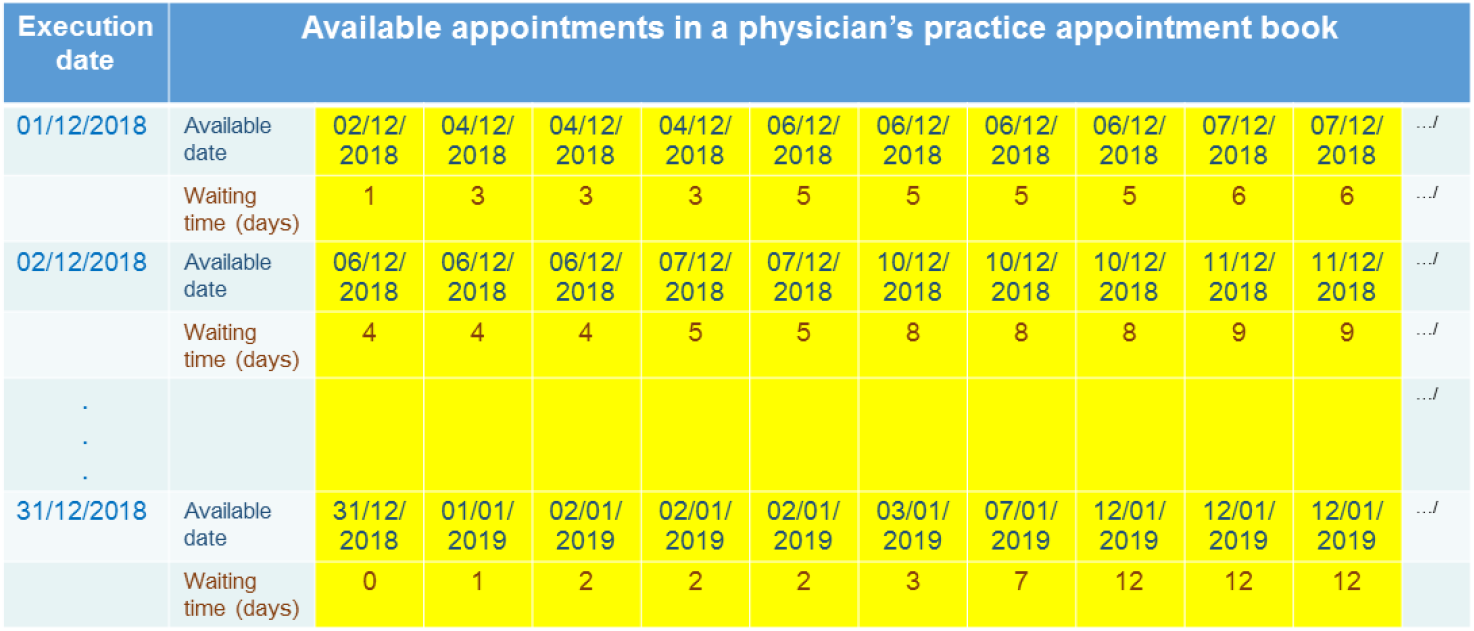
Schematic representation of WT calculation for three selected execution dates extracted from one physician practice appointment book.

The second step of the algorithm is to determine how many available appointments, offered to HMO members, can be utilized in the OWT calculation.

For the “specific” physician scenario, the daily demand is calculated by dividing the total number of actual visits of the physician by the number of days in the period (n=31). Under the steady state assumption (demand=supply), the number of the first available appointments that should be utilized for the calculation from the physician’s appointment book each day (supply), is equal to the daily demand. For “any” physician scenario, demand is calculated as the number of actual visits of all the physicians of the selected specialty in the region, divided by the number of days in the period (n=31). The supply, or the number of the first available appointments that should be utilized, from <u>all</u> appointment books each day, in each geographical region (supply), is equal to the above calculated demand. In both scenarios, calculation was done separately for each specialty.

Figure 2 demonstrates the process of determining supply and demand sizes in OWT calculation, in an illustrative region with two specialists. It is demonstrated for both “specific” and “any” physician scenarios, on a single execution date (01/12/2018).

**Figure 2:**
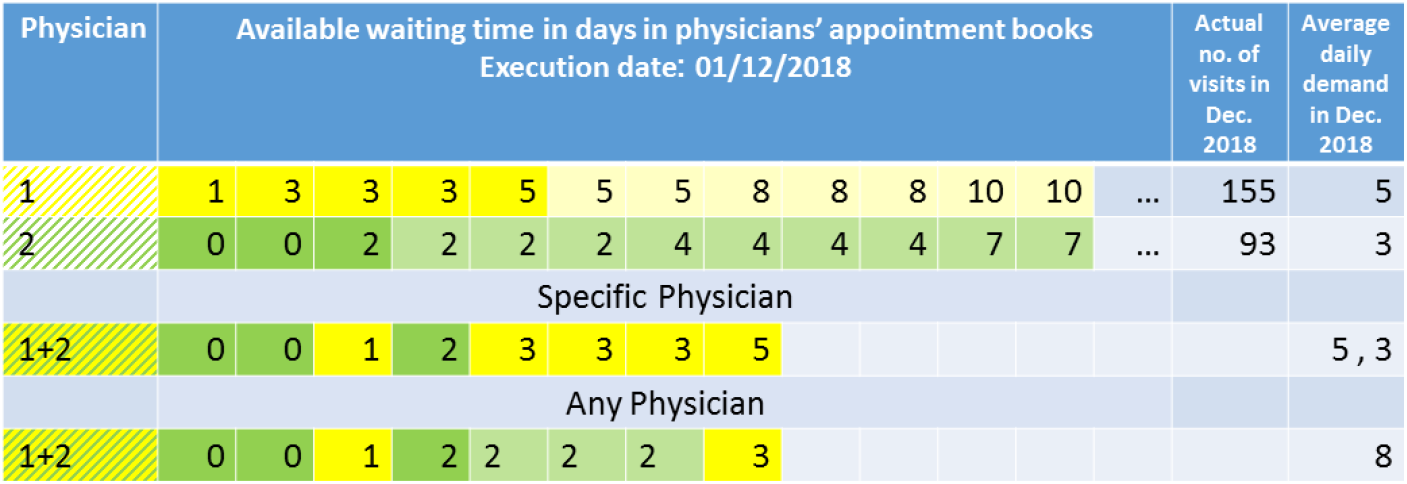
Scheme of supply and demand size determination, in “specific” vs “any” physician scenario, on a single execution date (01/12/2018). The yellow and green colors represent two illustrative physician practices (no. 1 no. 2). Dark shades of yellow and green represent the first available appointments equal to demand for each physician; light shades refer to all other available appointments for that physician.

The yellow and green colors represent two illustrative physician practices.

The number of the first available appointments taken from each appointment book was based on each physician’s average daily demand. In the “specific” physician scenario, the first five available appointments were extracted from the “yellow” physician (no. 1) and the first three available appointments were extracted from the “green” physician’s (no. 2) appointment book. In the “any” physician scenario, the number of the first available appointments that were utilized from appointment books was based on the average daily demand of all physicians in the region: In the illustration there are two physicians’ practices in the region, and the average daily demand for both physicians is eight. For this scenario only two of the soonest available appointments were extracted from the “yellow” physician while six of the soonest available appointments were extracted from the “green” physician’s appointment book.

The same process should be performed for each execution date. As seen in Figure 3, in the “specific” physician scenario five available appointments were extracted from the yellow physician and three were extracted from the green physician’s appointment book on each extraction date. In the “any” physician scenario the algorithm used eight appointments on each extraction date, but different numbers of available appointments were extracted from each physician’s practice appointment book on each date, considering the shortest waiting times that were calculated in the region.

**Figure 3:**
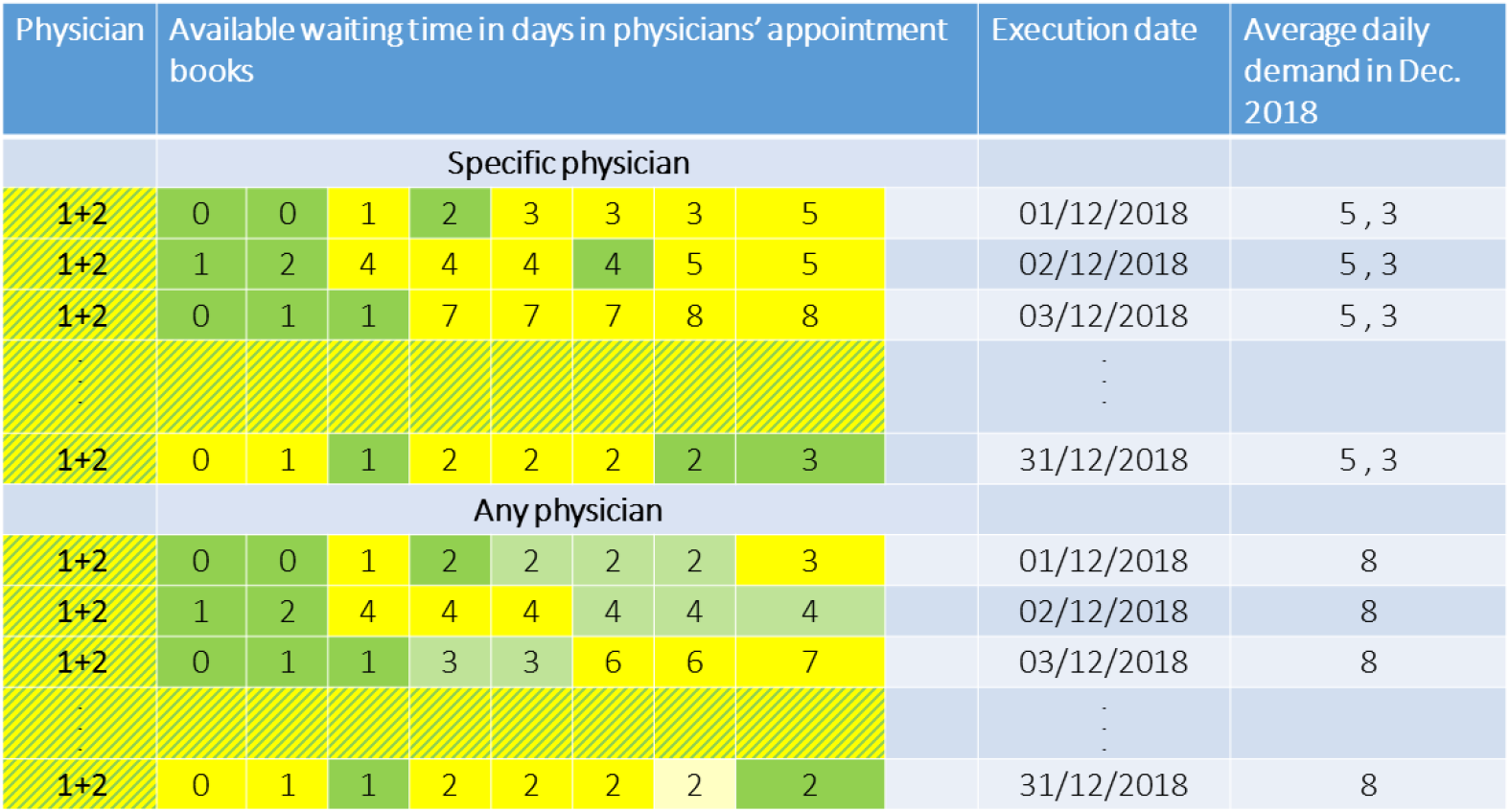
Extraction of available appointments, according to demand, in a determined specialty, region and period of time: “specific” vs “any” physician scenarios

The third phase of the algorithm is to calculate the distribution of OWT, expressed in days, for all execution dates for each specialty and geographical region. The histogram in Figure 4 demonstrates the distribution of OWT for an illustrative case of two physician practices in a given region. As expected, the right tale is longer and higher for “specific” physician than for “any” physician, since patients have to wait for longer periods in the case of a chosen physician. In the “any” physician scenario, more appointments are available sooner (shorter OWT), compared with specific physician.

**Figure 4:**
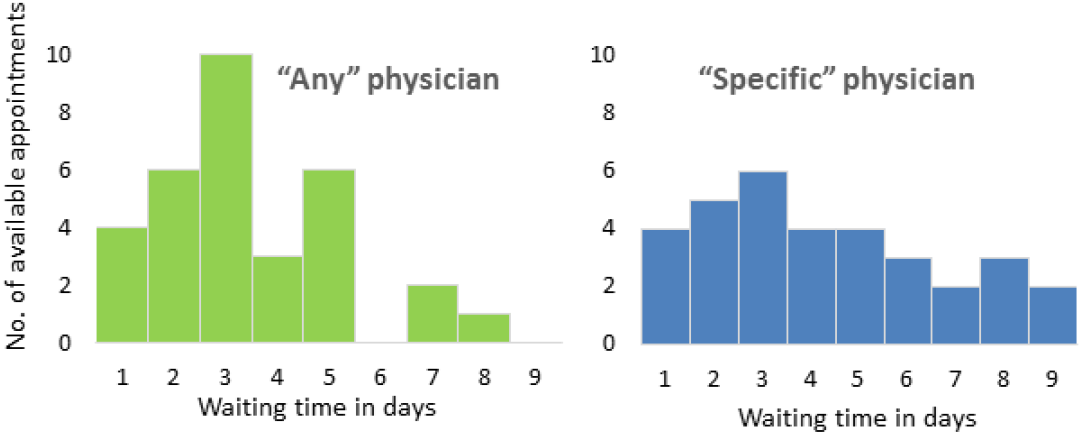
OWT histograms for a given specialty and region, in “any” vs. “specific” physician scenarios

The last phase of the algorithm is to calculate OWT distribution for geographical areas. In order to do so, OWT distributions of all “natural” areas, were summarized both at the district and at the national level. Distributions at each geographic level were used to calculate diverse statistical indices.

## Results

1,017,478 available appointments were collected during December 2018 from all computerized appointment books of 6,040 physician practices, covering five medical specialties, from all Israeli HMOs.

At the national level, median OWT for “specific” physician scenario ranged from 6 and 7 days, in ophthalmology and otolaryngology respectively, to 13 days in dermatology. Median OWT for “any” physician scenario ranged from 4 days, in gynecology and ophthalmology, to 8 days in dermatology (Figure 5). Median OWT in the “any” physician scenario were between 28% (otolaryngology) to 50% (gynecology) shorter compared to the “specific” physician scenario.

**Figure 5:**
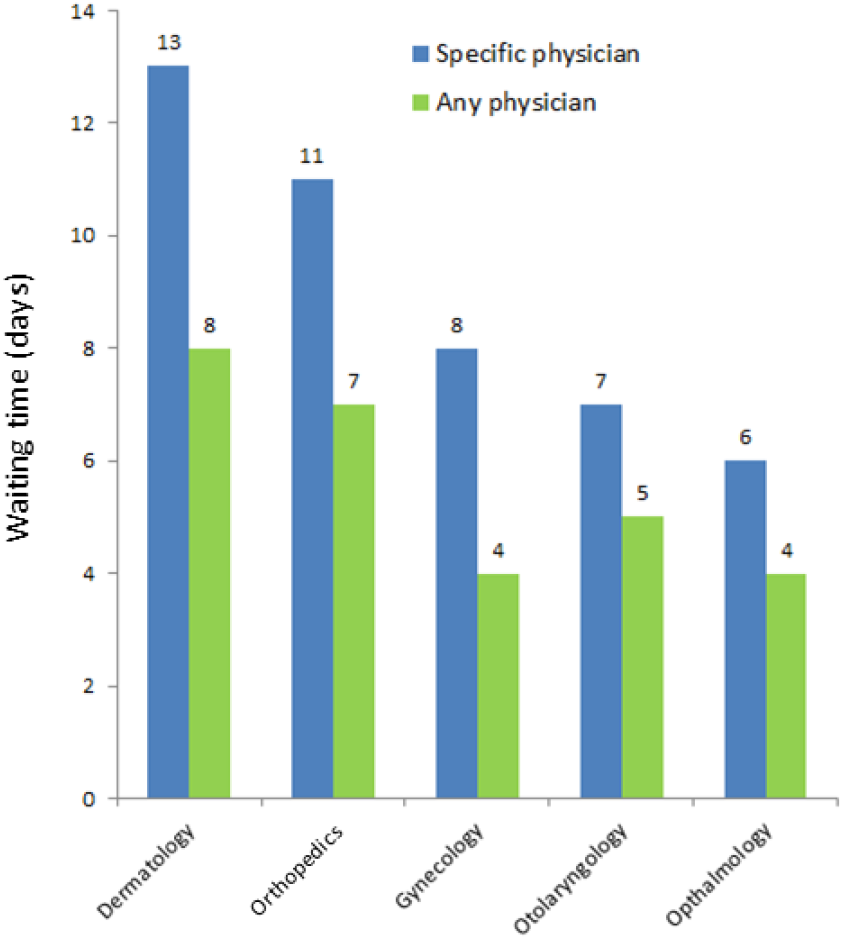
National level median OWT, by medical specialty: “specific” vs. “any” physician scenario

Mean OWT at the national level for “specific” physician ranged from 13.3 days (otolaryngology) to 21.7 days (obstetrics & gynecology). For “any” physician, mean OWT ranged from 4.9 (gynecology) to 9.5 days (dermatology). As expected, mean OWT are longer, compared with median OWT, since the mean takes into account a “long tail” that includes appointments with experts in high demand, for whom patients are willing to wait longer.

Israel is divided into 6 districts. Analysis of OWT by district demonstrated considerable geographical variation, with longest waiting times in the Southern, compared with the other districts, in 4 out of 5 specialties examined. For example, median OWT for consultation with an ophthalmologist, was more than three times longer in the Southern district, compared with Tel Aviv district (13.0 versus 4.0 days, respectively). The 75^th^ percentile of OWT for an ophthalmologist was 33 days in the Southern district, compared with 11 days in Tel Aviv. In Be’er-Sheva, the “capital” of the Southern district, the 75^th^ percentile of OWT for a specific physician, was 55 days, i.e. one in four consumers had to wait for more than 55 days to see an eye doctor for non-urgent care. Analysis of OWT at all geographical levels, from national level to city level, presents shorter OWT for “any” physician scenario than “specific” physician scenario, across the board. For example, in the city of Haifa, median wait for any physician was 3 days, (75^th^ percentile of 4 days), and for a specific physician was 4 days, (75^th^ percentile of 11 days). Other cities showed greater differences such as Jerusalem where the median wait for any physician was 3 days, compared to 8 days for specific physician.

## Discussion

This study presents the step-by-step development of a methodology that creates, for the first time, the basis for a national measurement of offered wait time (OWT) for community-based specialty care. The method is based on a common denominator that utilizes the existing CASS of the four healthcare providers. It provides information of great importance to patients, caregivers and policy makers, enabling transparency within the healthcare system and providing the opportunity to improve access to care.

Long waiting times may reflect socioeconomic inequalities. Moscelli et al demonstrated socioeconomic inequalities in WT for non-emergency coronary revascularization in the British National Health Service, a publicly funded health system. Only a fraction of the inequalities were explained by patient choice.[20] Siciliani and Verzulli found that more educated patients had lower waiting times for specialist consultation and nonemergency surgery.[9] In Israel, the expected waiting time for surgeries was longer for patients with more comorbidities and those in the geographic periphery.[21] Long WT can also increase socioeconomic inequalities, when the weakest segments of the population find it more difficult to find private or other pathways to shorten WT.[22] In a representative survey carried out in Israel during 2018, 32% of respondents belonging to the lower quintile of income reported forgoing some kind of medical care during the year prior to the survey due to long WT. [17]

Our study demonstrated considerable variations in OWT between geographic regions, for example much longer OWT for consultation with an ophthalmologist in the Southern district, compared with Tel Aviv district. These differences, along with the difference in OWT across cities, highlight the issues of equity and the need for a transparent reporting of these disparities to decision makers and the public.

The methodology presented in this study should be compared with existing WT metrics that utilize a prospective approach. The approach of calculating WT based on the first available appointment or the third next available appointment can cause bias, since it is sensitive to last minute cancellation and other unexpected events. Both the first or third available appointments might, therefore, under-estimate OWT. On the other hand, fourth or fifth available appointment can over-estimate OWT, since less than 4 patients were looking for available appointments each day in a third of the physician practices in our study. Our approach uses all first available appointments for each physician (supply), and utilizes for calculation an amount equal to the average daily number of visits (demand). Since no physician had daily average demand greater than 50 appointments, it was deemed that this approach produced the best estimation of real OWT distribution. The number of available appointments introduced into the algorithm can be customized to different health systems.

When comparing OWT with AWT, two situations may act in opposing directions: people do not necessarily choose the first available offered appointment (as calculated in OWT measurement), and may prefer a delayed appointment, due to time preferences, physician’s requests, follow-up visits etc. This situation can lengthen AWT compared with OWT. On the other hand, HMOs invest a lot of effort in order to shorten long OWT by managing computerized waiting lists, sending automated repeated reminders to confirm arrival or cancellation etc. These efforts can shorten AWT compared with OWT. Measurement of OWT is important in its own right, most accurately representing patients’ experience when trying to book a specialist appointment, and should be one of the basic elements for measuring healthcare systems’ performance.

Some assessments of waiting time rely on survey data, as has been previously conducted in Israel and elsewhere.[11, 15, 17] Comparison of WT estimated by the proposed methodology to WT reported by a consumer survey of a representative Israeli sample that took place in parallel months,[17] yielded similar results. This strengthens the contribution of the methodology presented in our study as a less expensive, easier to use alternative, compared with consumer surveys for the estimation of WT. While a well-organized survey may minimize biases in estimation of OWT, a complete administrative database produces better accuracy, at lower costs.

Another contribution of the methodology is its ability to estimate WT for two possible scenarios of physician choice - “specific” or “any” physician. Longer wait for a chosen doctor might cause patients to prefer “any” doctor, thus seeing a different doctor on each visit in order to reduce WT. There is a tradeoff between continuity of care and waiting time. High continuity of care is typically associated with better health outcomes but has also been associated with longer WT.[23] Few studies have addressed this difference. Fyie et al [24] reported that patients selecting a particular surgeon led to what they termed ‘voluntary delays’ in surgery or consultation, though this type of delay was not tracked in the system. A Canadian patient satisfaction survey reported that median wait time was longer for patients requesting a specific physician compared to any physician.[25] However, to the best of our knowledge, the ability to calculate waiting times, based on administrative data, separately for “specific” versus “any” physician, is unique.

Although developed in a given setting of one nation’s public healthcare system, the methodology described here is very flexible and can be adapted to diverse settings. When adapting the algorithm, each healthcare provider (or regulating authority) can decide on the reasonable distance that a patient can travel for a specialist, and widen or narrow the geographic units of reference, accordingly. The method can also be adopted to measure OWT for consultation with physicians from other medical specialties, as well as primary care; it can be used in public and private health systems. Its core competencies are derived from its ease of using existing administrative data, allowing ongoing monitoring at relatively low costs. Once the physician scenario and number of first available appointments are set, it can serve for ongoing monitoring of OWT at the local, regional and national levels. The complexity of comparing waiting times between countries was demonstrated by Viberg et al,[8] showing the considerable barriers caused by different measurement systems. The authors called for “a more coherent approach to waiting time measurement”. The methodology developed here has succeeded in overcoming system differences between HMOs to create a national measure, and may in the future lead the way for standardization and comparison across nations.

The algorithm presented in this study bears several limitations: 1) The assumption that the number of HMO enrollees seeking an available appointment is identical throughout all days of a given month. This number, representing the actual average daily demand, might change between weekdays and weekends, or be higher on the first workday of the week, for instance. This assumption can be resolved, if the information on volume by type of day is available, by calculating the average daily demand by type of day (weekday, weekend, vacation) and accounting for the weekly cycle and the effect of holidays;

2) In a steady state, demand equals supply, i.e. the number of HMO enrollees looking for an appointment equals the number of actual visits to a specialist practice in a given period of time. However, in the real world, there are two exceptions that should be considered in estimating OWT. Some patients visit the physician without scheduling an appointment (“walk-ins”). OWT for these patients is 0 days, because the physician accepted them on the same day. Under the steady state assumption, we included these visits in calculation of the demand, resulting in over-estimation of OWT. On the other hand, 19% of patients who needed a consultation with a specialist, did not book an appointment because of long WT.[26] Thus, the real demand may be higher than the actual number of visits. This leads to under-estimation of OWT. If the proportion of patients who give up in advance is known, one can correct the demand side of the OWT estimation accordingly. Our study did correct the demand for such situations, which presumably balance each other out;

3) Extraction of data took place once a day, not allowing for cancelations and rescheduling of appointments after extraction time to be included in the calculation.

The ideal measurement of WT may be obtained by developing a computerized process that continuously collects the following parameters: The time when a patient enters the system, a list of available appointments for the relevant physicians at this moment and the actual appointment that the patient chooses. These measures will be incorporated into the next stage of the national measurement.

## Conclusions

The novelty of this methodology lies in the utilization of existing computerized scheduling systems to create a national measurement of WT, and the integration of patient preferences for physician choice, which allows analysis of the tradeoff between continuity of care and waiting time. The designed method was further able to overcome differences in IT systems between health providers, thus providing a tool for the comprehensive assessment of waiting times for specialist care, and supplying essential information to policy makers and the public. This is a vast improvement on previously available survey data and anecdotal media reporting of WT, which was the case in Israel. This low-cost method allows ongoing monitoring and periodic public reporting of data. It is already being utilized by the MOH as an ongoing tool for monitoring and dialogue with the HMOs and is reported to the public on a quarterly basis.

This tool can inform better allocation of resources, both by HMO managers and by the regulator. Future comparative reporting of WT by health provider will provide transparency and allow patients to make informed choices. Accurate assessment of WT by geographic regions can identify disparities in care access and help target interventions. These might reduce WT and improve equity and ultimately strengthen the public healthcare system.

## Data Availability

Data is available upon request

